# The “Weekend Warrior” Physical Activity Pattern and Cardiometabolic Health: A Pooled Harmonized Individual Participant Analysis of the ProPASS Consortium

**DOI:** 10.64898/2026.07.21.26358626

**Authors:** Shan Jiang, Matthew N Ahmadi, Nicholas A. Koemel, Amy S Ha, Raaj Kishore Biswas, Joanna M Blodgett, John J Mitchell, Borja del Pozo Cruz, Richard M. Pulsford, Kristin Suorsa, Dick H.J. Thijssen, Esmee A Bakker, Carlos Celis-Morales, Peter J. Johansson, Pasan Hettiarachchi, Sari Stenholm, Gita Mishra, Sarah Kozey Keadle, Vegar Rangul, Nidhi Gupta, Stavros Kyriakidis, Annemarie Koster, Marvin Y. Chong, Andrew J Atkin, I-Min Lee, Mark Hamer, Emmanuel Stamatakis, the ProPASS Collaborators, Hans Savelberg, Bastiaan de Galan, Carla van de Kallen, Thijs M.H. Eijsvogels

**Affiliations:** Department of Health and Physical Education, The Education University of Hong Kong, Hong Kong SAR, China; Turner Institute for Brain and Mental Health, School of Psychological Sciences, Faculty of Medicine, Nursing and Health Sciences, Monash University, Melbourne, Victoria, Australia; Mackenzie Wearables Research Hub, Charles Perkins Centre, The University of Sydney, Sydney, New South Wales, Australia; Victorian Heart Institute, Monash University, Melbourne, Victoria, Australia; Department of Sports Science and Physical Education, The Chinese University of Hong Kong, Hong Kong SAR, China; Charles Perkins Centre, School of Health Sciences, Faculty of Medicine and Health, The University of Sydney, Sydney, New South Wales, Australia; Snow Vision Accelerator, Save Sight Institute, Faculty of Medicine and Health, The University of Sydney, Sydney, New South Wales, Australia; Institute of Sport, Exercise and Health, Division of Surgery and Interventional Sciences, Faculty of Medical Sciences, University College London, 170 Tottenham Court Road, London, W1T 7HA, UK; University College London Hospitals, National Institute for Health and Care Research Biomedical Research Centre, London, United Kingdom; Department of Sport Science, Faculty of Medicine, Health, and Sports, Universidad Europea de Madrid, Campus de Villaviciosa de Odón, Madrid, Spain; University of Exeter, Faculty of Health and Life Sciences, Exeter, UK; Department of Public Health, University of Turku and Turku University Hospital, Turku, Finland; Centre for Population Health Research, University of Turku and Turku University Hospital, Finland; Radboud University Medical Center, Department of Medical BioSciences, Nijmegen, The Netherlands; Research Institute for Sport and Exercise Sciences, Liverpool John Moores University, Liverpool, United Kingdom; Department of Medical BioSciences, Radboud university medical center, Nijmegen, The Netherlands; Department of Medical BioSciences, Radboud University Medical Center, route 928, Nijmegen, The Netherlands; University of Glasgow, School of Cardiovascular and Metabolic Health, Glasgow, UK; Universidad Católica del Maule, Human Performance Lab, Education, Physical Activity, and Health Research Unit, Talca, Chile; High-Altitude Medicine Research Centre (CEIMA), Universidad Arturo Prat, Iquique, Chile; Department of Occupational and Environmental Medicine, Uppsala University Hospital, Uppsala, Sweden; Department of Medical Sciences, Occupational and Environmental Medicine, Uppsala University, Uppsala, Sweden; Research Services, Turku University Hospital and University of Turku, Finland; The University of Queensland, School of Public Health, Brisbane, Australia; Department of Kinesiology and Public Health, California Polytechnic State University, San Luis Obispo, CA, USA; HUNT Research Centre, Department of Public Health and Nursing, Faculty of Medicine and Health Sciences, Norwegian University of Science and Technology (NTNU), Levanger, Norway; The National Research Centre for the Working Environment, Department of Ergonomics and Musculoskeletal Health, Copenhagen, Denmark; Department of Social Medicine, CAPHRI Care and Public Health Research Institute, Maastricht University, Maastricht, The Netherlands; Department of Social Medicine, CARIM Cardiovascular Research Institute Maastricht, Maastricht University, Maastricht, the Netherlands; School of Health Sciences, University of East Anglia, Norwich, UK; Brigham and Women’s Hospital, Harvard Medical School, Division of Preventive Medicine, Boston, MA, USA; Department of Epidemiology, Harvard TH Chan School of Public Health, Boston, MA, USA; Department of Human Biology and Movement Sciences, NUTRIM School of Nutrition and Translational Research in Metabolism, Faculty of Health, Medicine and Life Sciences, Maastricht University, Maastricht, The Netherlands; CARIM Cardiovascular Research Institute Maastricht, Maastricht University, Maastricht, the Netherlands; Department of Internal Medicine, Maastricht University Medical Center+ (MUMC+), Maastricht, the Netherlands; Department of Internal Medicine, Radboud University Medical Center, Nijmegen, the Netherlands

**Author notes:** Emmanuel Stamatakis is the corresponding author. Mark Hamer and aEmmanuel Stamatakis are joint senior authors. Shan Jiang and #Matthew N Ahmadi contributed equally.

**Keywords:** weekend warrior, physical activity, accelerometry, cardiometabolic health

## Abstract

**Background:** While “weekend warrior” (WW) physical activity (PA) pattern has been associated with cardiovascular benefits, most existing evidence is based on self-reported PA. Evidence using harmonized, thigh-worn accelerometry to examine associations between the weekend warrior phenotype and comprehensive cardiometabolic profiles remains limited.

**Methods:** We pooled harmonized individual-participant data from six cohorts in the Prospective Physical Activity, Sitting and Sleep (ProPASS) Consortium. In this cross-sectional analysis, participants were classified as inactive (<150 min/week MVPA), weekend warriors (≥150 min/week with ≥50% accumulated on 1-2 days), or regularly active ≥150 min/week but not meeting the WW criterion). A composite cardiometabolic score averaged eight standardized indicators: triglycerides, HDL cholesterol, total cholesterol, HbA1c, body mass index, waist circumference, systolic blood pressure, and diastolic blood pressure. Generalized linear models were used to estimate associations between PA patterns and both composite and individual cardiometabolic outcomes, adjusted for age, sex, smoking, alcohol intake, self-rated health, CVD history, medication use, blood biomarker fasting status, and cohort.

**Results:** Among 13,904 adults (mean age 54.3±9.5 years; 54.7% women), 61.8% were inactive, 24.5% weekend warriors, and 13.7% regularly active. Compared with inactivity, both active patterns were associated with more favorable composite cardiometabolic scores (WW: β = -0.118 (95% CI: -0.142, -0.095); regularly active: β = -0.111 (95% CI: -0.141, -0.081)), with negative values indicating better cardiometabolic health. Weekend warriors and regularly active adults showed broadly similar associations across individual markers, including lower adiposity (BMI and waist circumference) and more favorable metabolic biomarkers (higher HDL cholesterol; lower triglycerides and HbA1c), with no meaningful differences between the two active patterns for the composite score or any individual outcome. Associations with total cholesterol and blood pressure were small.

**Conclusions:** Both active patterns were associated with more favorable cardiometabolic profiles than inactivity, and profiles were broadly comparable whether MVPA was concentrated on 1-2 most active days (weekend warrior) or accumulated more regularly across the week.

## Introduction

Cardiometabolic diseases are a leading global cause of morbidity and premature mortality.^1,2^ In clinical and public health practice, cardiometabolic health is routinely assessed using a cluster of metabolic, adiposity, and hemodynamic markers for risk stratification and preventive interventions.^3^

Physical activity (PA) is widely recognized as beneficial for health and is consistently associated with reduced risks of cardiovascular disease and mortality.^4–6^ Current guidelines, including those by the World Health Organization (WHO) and American Heart Association, recommend that adults aged 18 to 64 years accumulate at least 150 minutes of moderate-to-vigorous physical activity (MVPA) per week but do not specify how this activity should be distributed across the week.^1,7^ In contrast, the UK National Health Service advises that MVPA be spread “evenly over 4 to 5 days per week or every day”.^8^ However, whether concentrating MVPA into 1-2 days yields cardiometabolic health profiles comparable to more evenly distributed MVPA remains unclear.

One commonly studied alternative is the “weekend warrior” (WW) pattern, a concentrated across-day MVPA pattern that is not necessarily restricted to weekends or consecutive days.^9^ WW is typically defined as achieving guideline-consistent MVPA while accruing ≥50% of weekly MVPA on the 1-2 most active days; “regular” activity denotes meeting the same weekly MVPA volume without this degree of concentration. Recent evidence suggests that the WW pattern may be associated with lower risks of incident disease and mortality.^6,9–13^ For example, in a prospective cohort of approximately 350,000 individuals, WW and regularly active patterns showed similarly lower risks of cardiovascular mortality,^14^ and analyses of 89,573 UK Biobank participants found that achieving guideline-consistent PA volumes reduced the risk of more than 200 diseases, particularly cardiometabolic conditions, with comparable benefits for WW and regularly active adults.^15^ However, most evidence is based on self-reported PA data, which is prone to recall and social-desirability biases,^14,16^ and comes from single cohorts in relatively homogeneous settings, limiting generalizability.^10,11,15,17,18^

To enhance generalizability, evidence is needed beyond single cohorts and self-reported PA. Individual participant data (IPD) analyses of harmonized multi-cohort data can improve statistical power and precision and allow consistent definitions of PA exposures.^19,20^ Thigh-worn accelerometers provide device-measured information on PA volume, intensity, and temporal patterning across the week,^19,21–23^ and may reduce misclassification compared with self-reported PA. Building on these methodological advances, large pooled IPD resources of device-measured PA now enable the evaluation of WW and regularly active patterns across diverse international populations.^24^

Using data from six cohorts within the Prospective Physical Activity, Sitting and Sleep (ProPASS) Consortium, we conducted a harmonized pooled IPD analysis to examine whether a concentrated MVPA pattern (“weekend warrior”) is associated with cardiometabolic health comparable to a less concentrated (regular) pattern. We assessed a composite cardiometabolic health score and its component metabolic, anthropometric, and hemodynamic measures. As a secondary aim, we examined dose-response associations between weekly MVPA and cardiometabolic outcomes across activity patterns.

## Methods

### Consortium Sample and Harmonization Process

The Prospective Physical Activity, Sitting, and Sleep (ProPASS) Consortium is an international research platform that integrates existing and future observational studies that use wearable devices to assess movement behaviors.^22^ For this study, cross-sectional data were pooled from 6 cohorts across 5 countries, each using standardized 7-day, 24-hour thigh-worn accelerometry protocols, yielding a harmonized dataset of thigh-worn accelerometry data as described previously.^21^ Participants in the study were drawn from the pooled ProPASS data resource comprising 6 participating studies, including ActivPAL3/4 accelerometers (the 1970 British Birth Cohort Study [BCS70],^25^ The Maastricht Study [TMS],^26^ the Australian Longitudinal Study on Women’s Health [ALSWH],^27^ and the Nijmegen Exercise Study [NES]),^27^ Axivity devices (the Finnish Retirement and Aging Study [FIREA]),^28^ and ActiGraph devices (the Danish Physical Activity Cohort with Objective measurements [DPhacto]).^29^ Ethics approvals and participant consent were obtained independently at the cohort level, and all covariates, outcomes, and raw accelerometer data were harmonized and pooled into a single dataset at the University of Sydney. Detailed descriptions of each study are available in Table S1 and have been published previously.^21^ Device models/deployment details in Table S2.

### Physical Activity Patterns

The raw accelerometer data were processed centrally using ActiPASS v1.56, a validated, device-agnostic platform specifically designed for thigh-worn accelerometers, compatible with all device brands included in this analysis.^30–34^ ActiPASS identifies behaviors in 2 second (s) windows with a 50% overlap, resulting in a resolution of 1 s epochs, and implements non-wear,^35^ sleep detection,^33^ and intensity classification based on activity type and stepping cadence.^34,36–38^ Five movement behaviors were classified: sleep, sedentary behavior (SB) (sitting or lying episodes outside of sleep intervals), standing, light-intensity physical activity (LIPA) (ambulatory movement without purposeful walking, and walking with cadence <100 steps/min), and MVPA (running, cycling, inclined stepping, walking with cadence ≥100 steps/min). MVPA was classified using 1-minute cadence windows; therefore, MVPA estimates may differ from prior ProPASS studies using different epoch lengths and/or intensity derivation methods.^36,39^ Weekly-equivalent MVPA (min/week) was calculated by standardizing MVPA accrued on valid wear days to a 7-day week (i.e., averaging across valid days and multiplying by 7) and used to classify participants as active (≥150 min/week) versus inactive (<150 min/week). Among active participants, WW (concentrated) were defined as accruing ≥50% of weekly MVPA on the two most active days; active participants who did not meet this criterion were classified as regularly active. The two most active days were not required to be weekend or consecutive, and WW was used as a label for concentration rather than weekend-only activity. Participants with ≥4 valid days (≥20 h/day), including ≥1 weekend day, were included. Days were defined from 00:00 to 23:59 (local time).

### Cardiometabolic outcomes

#### Composite Cardiometabolic Health Score

Adiposity indicators, including body mass index (BMI; kg/m²) and waist circumference (cm), and blood pressure (BP) were measured by trained staff following standardized clinic or home procedures (see Table S3). BP was recorded using automated monitors after a 5-10 minute seated rest, and the mean systolic (SBP) and diastolic (DBP) values were calculated from two or three readings, depending on cohort.^21^ Blood-based markers were available in five cohorts (excluding DPhacto) and included HDL cholesterol (mmol/L), total cholesterol, triglycerides (mmol/L), and HbA1c (mmol/mol; measured only in ALSWH, BCS70, and TMS). Detailed assay methods, quality control procedures, and coefficients of variation are presented in Table S3 and have been described previously.^19,21,33^

Individual cardiometabolic indicators were standardized to z-scores using the mean and SD of the pooled analytic sample (across cohorts) and averaged to generate a composite cardiometabolic health score. The score comprised eight indicators: BMI, waist circumference, SBP, DBP, triglycerides, HDL cholesterol, total cholesterol, and HbA1c. HDL cholesterol was inversely coded (multiplied by -1) so that higher values consistently indicated worse cardiometabolic health across all components. Higher composite scores indicate worse cardiometabolic health. The composite score was calculated only among participants with complete data on all eight indicators; because HbA1c was available only in ALSWH, BCS70, and TMS, the composite analyses mainly reflect these cohorts and exclude DPhacto.

### Covariates

For each participating cohort, covariates were assessed during clinic or home visits. Harmonized covariates were selected based on data availability and their associations with PA and cardiometabolic health outcomes.^19,23,40,41^ These covariates across all cohorts included age (in years), sex (male/female), smoking status (non-smoker/current smoker), alcohol consumption (categorized into cohort-specific tertiles based on weekly intake), self-rated health (measured on a 5-point Likert scale), self-reported medication use (blood pressure, glucose, and lipid-lowering medications), self-reported history of CVD, and cohort.^19,21,23^ Fasting status was included as a covariate in analyses of blood biomarker outcomes.^19^ A subset of cohorts contributed additional information on education (n = 4 cohorts; categorized as none or less than high school, high school qualification, further education qualification, and university degree or higher), occupational class (n = 5; not working, low, intermediate, or high occupational class), diet (n = 3; low, low-moderate, moderate-high, or high fruit and vegetable consumption), and mobility limitations (n = 4; questionnaire scores ranging from 0 to 100).^3^ Details of the covariate harmonization procedures are provided in Table S4.

### Statistical Analyses

We conducted a one-stage individual participant data meta-analysis using generalized linear regression with a Gaussian distribution to estimate the associations between different PA patterns and composite cardiometabolic health (z-score), as well as individual cardiometabolic health outcomes.^42^ All statistical analyses were conducted in RStudio (version 4.2.3). Results are presented as beta coefficients with 95% confidence intervals (CIs), and assumptions for regression analyses were assessed using residual plots and leverage versus residual squared plots to confirm model validity. We applied three regression models: Model 1 was adjusted for demographic variables (age, sex, and cohort), and Model 2 further adjusted for alcohol consumption, smoking status, and fasting status (for blood biomarker outcomes). Model 3 additionally included self-rated health, history of CVD, and self-reported use of medications for blood pressure, glucose, and lipid management.^19,21,23^ We examined dose-response relationships between weekly MVPA and cardiometabolic health measures by modelling MVPA as a continuous exposure using natural cubic splines (df = 4) and including interactions between the spline terms and PA pattern (inactive, weekend warrior, regularly active) to allow the MVPA-outcome association to vary by pattern. We reported adjusted curves with 95% CIs, using 150 min/week as the reference, and restricted plots to the 1^st^-99th percentile of MVPA within each pattern group to minimize extrapolation. Evidence of non-linearity was assessed by comparing the spline model with a corresponding linear MVPA model using nested-model F tests.

To assess the robustness of our findings, we conducted additional analyses: (1) using an alternative WW definition (≥150 min/week MVPA with ≥75% accumulated over 1-2 days);^15^ (2) reclassifying PA patterns using the sample median MVPA (≥117.1 min/week) as the weekly MVPA cut-off for group assignment (WW/regular active vs inactive); (3) restricting the analytic sample to participants meeting stricter wear-time criteria (≥5 valid wear days [≥20 h/day] and ≥1 weekend day observed); and (4) accounting for MVPA volume by (i) additionally adjusting for total weekly MVPA (minutes/week) in a separate Model 3-based analysis and (ii) conducting an exploratory volume-restricted analysis (WW/regular: 150-300 min/week; inactive: <150 min/week) and re-estimating Model 3 associations for SBP and DBP.

Sensitivity analyses were performed using data available from the ALSWH, BCS70, DPhacto, and TMS cohorts, which included additional variables on socioeconomic status (education), occupational class, diet (fruit and vegetable consumption), self-rated health and mobility limitations. In primary models, we adjusted for self-reported use of medications for blood pressure, glucose, and lipid management. As a sensitivity analysis for blood pressure outcomes, SBP and DBP values were additionally adjusted by adding 10 mm Hg for participants using antihypertensive medication, in accordance with established protocols.^44,45^ The medication use of antihypertensive was recorded across all cohorts. An additional sensitivity analysis was conducted after excluding participants with prevalent CVD (e.g., heart disease, myocardial infarction, angina, stroke; see Table S4 for cohort-specific definitions) or medication use.^3^ This pooled, harmonized cross-sectional analysis is reported in accordance with the STROBE guidelines^43^; the completed STROBE checklist is provided in the Supplemental Methods S2.

## Results

### Sample Characteristics

We included 13,904 participants (mean [SD] age, 54.3 [9.5] years; 54.7% female) with data for ≥1 cardiometabolic outcome. In total, 13,915 participants met accelerometer inclusion criteria (≥4 valid wear days, including ≥1 weekend day). Valid wear days were 4 (4.6%), 5 (6.0%), 6 (28.3%), and 7 (61.1%); median 7 (IQR 6-7). The composite-score analyses included 9,866 participants with complete data on all eight-component metrics. (Table 1; Figures S1-S2). HbA1c was available only in ALSWH, BCS70, and TMS, and blood biomarkers were not available in DPhacto.

**Table 1.** Sample characteristics by physical activity pattern (inactive, weekend warrior, regularly active)

| Characteristic | Total | Regularly Active | Weekend Warrior | Inactive |
| --- | --- | --- | --- | --- |
| Sample | 13904 | 1909 (13.7%) | 3407 (24.5%) | 8588 (61.8%) |
| Age, mean (SD), years | 54.3 (9.5) | 53.2 (9.0) | 55.0 (9.4) | 54.3 (9.7) |
| Female, n (%) | 7608 (54.7) | 1068 (55.9) | 1973 (57.9) | 4567 (53.2) |
| LIPA min/week (median [IQR]) | 1014 (793.3, 1264.0) | 1095 (866, 1326) | 1041 (834, 1279) | 987 (768, 1241) |
| MPA, min/week (median [IQR]) | 105 (56, 175) | 229 (176, 314) | 190 (156, 250) | 65 (37.8, 100) |
| VPA, min/week (median [IQR]) | 1 (0, 12) | 9 (0, 46) | 10 (0, 50) | 0 (0, 3) |
| MVPA, min/week (median [IQR]) | 117.1 (60, 199.5) | 258 (199, 356) | 217 (178, 286) | 72 (40, 108) |
| Sleep, min/day, (median [IQR]) | 430.8 (391, 471.3) | 428 (389, 464) | 429 (392, 467) | 432 (391, 474) |
| Current smoker, n (%) | 1956 (14.14) | 200 (10.54) | 297 (8.74) | 1459 (17.10) |
| Self-rated health, n (%) |  |  |  |  |
| Excellent | 1630 (11.93) | 327 (17.55) | 531 (15.88) | 722 (8.59) |
| Very good | 4365 (31.95) | 672 (36.07) | 1195 (35.74) | 2498 (29.71) |
| Good | 5901 (43.19) | 699 (37.52) | 1364 (40.79) | 3838 (45.65) |
| Fair | 1516 (11.09) | 144 (7.73) | 234 (7.0) | 1138 (13.54) |
| Poor | 252 (1.84) | 21 (1.13) | 20 (0.60) | 211 (2.51) |
| Alcohol consumption, n (%) |  |  |  |  |
| Tertile 1 (lowest) | 4085 (33.94) | 547 (33.21) | 902 (30.02) | 2636 (35.70) |
| Tertile 2 | 4085 (33.94) | 581 (35.28) | 1080 (35.94) | 2424 (32.83) |
| Tertile 3 (highest) | 3865 (32.11) | 519 (31.51) | 1023 (34.04) | 2323 (31.46) |
| Medication use, n (%) | 4072 (30.9) | 451 (24.6) | 819 (26.2) | 2802 (34.1) |
| Prevalent CVD, n (%) | 1392 (10.1) | 157 (8.28) | 288 (8.49) | 947 (11.1) |
| Cardiometabolic markers |  |  |  |  |
| BMI, kg/m<sup>2</sup>, mean (SD) | 26.8 (4.47) | 25.8 (4.0) | 25.8 (3.7) | 27.5 (4.7) |
| Waist circumference, cm, mean (SD) | 93.8 (13.1) | 90.6 (12.5) | 90.5 (11.8) | 95.7 (13.4) |
| Men | 99.8 (11.2) | 97.4 (10.4) | 97.1 (10.0) | 101.0 (11.4) |
| Women | 88.9 (12.6) | 85.4 (11.4) | 85.8 (10.7) | 91.5 (13.2) |
| Total cholesterol, mmol/L, mean (SD) | 5.26 (1.01) | 5.3 (1.0) | 5.33 (0.99) | 5.23 (1.02) |
| HDL cholesterol, mmol/L, mean (SD) | 1.55 (0.43) | 1.62 (0.44) | 1.64 (0.44) | 1.5 (0.42) |
| Triglycerides, mmol/L, mean (SD) | 1.44 (0.78) | 1.34 (0.75) | 1.31 (0.72) | 1.51 (0.8) |
| HbA1c |  |  |  |  |
| mmol/mol, mean (SD) | 37.7 (7.01) | 36.7 (6.03) | 36.7 (5.83) | 38.4 (7.52) |
| % | 5.6 (0.64) | 5.5 (0.55) | 5.51 (0.53) | 5.66 (0.7) |
| Systolic blood pressure, mmHg*, mean (SD) | 129.77 (16.62) | 128.59 (16.84) | 129.01 (16.71) | 130.33 (16.52) |
| Diastolic blood pressure, mmHg*, mean (SD) | 76.45 (9.83) | 76.31 (9.71) | 76.0 (9.77) | 76.66 (9.88) |

LIPA, light-intensity physical activity; MPA, moderate intensity physical activity.
VPA, vigorous intensity physical activity; MVPA, moderate-to-vigorous intensity physical activity
HDL, high-density lipoprotein
WW = weekend warrior; defined as ≥150 minutes/week of MVPA with ≥50% of total weekly MVPA accumulated on 1–2 days.

When stratified at guideline-recommended thresholds (≥150 minutes of MVPA per week), 8588 participants (61.8%) were classified as inactive, 3407 (24.5%) as WW (ie, ≥50% of total MVPA accumulated within 1-2 days), and 1909 (13.7%) as regularly active ((i.e., meeting ≥150 min/week but not meeting the WW criterion)). Median total weekly MVPA was highest in the regularly active group (258 minutes [IQR, 199-356 minutes]), followed by the weekend warrior group (217 minutes [IQR, 178-286 minutes]), and the inactive group (72 minutes [IQR, 40-108 minutes]) (Table 1 and Figure 1). Most participants were non-smokers (85.4%), reported good or better self-rated health (87.1%), were not taking lipid-lowering, antihypertensive, or glucose-lowering medications (69.1%), and had no history of cardiovascular disease (89.9%). Sample characteristics using alternative definitions of the WW pattern are provided in Tables S5 and S6.

**Figure 1.**
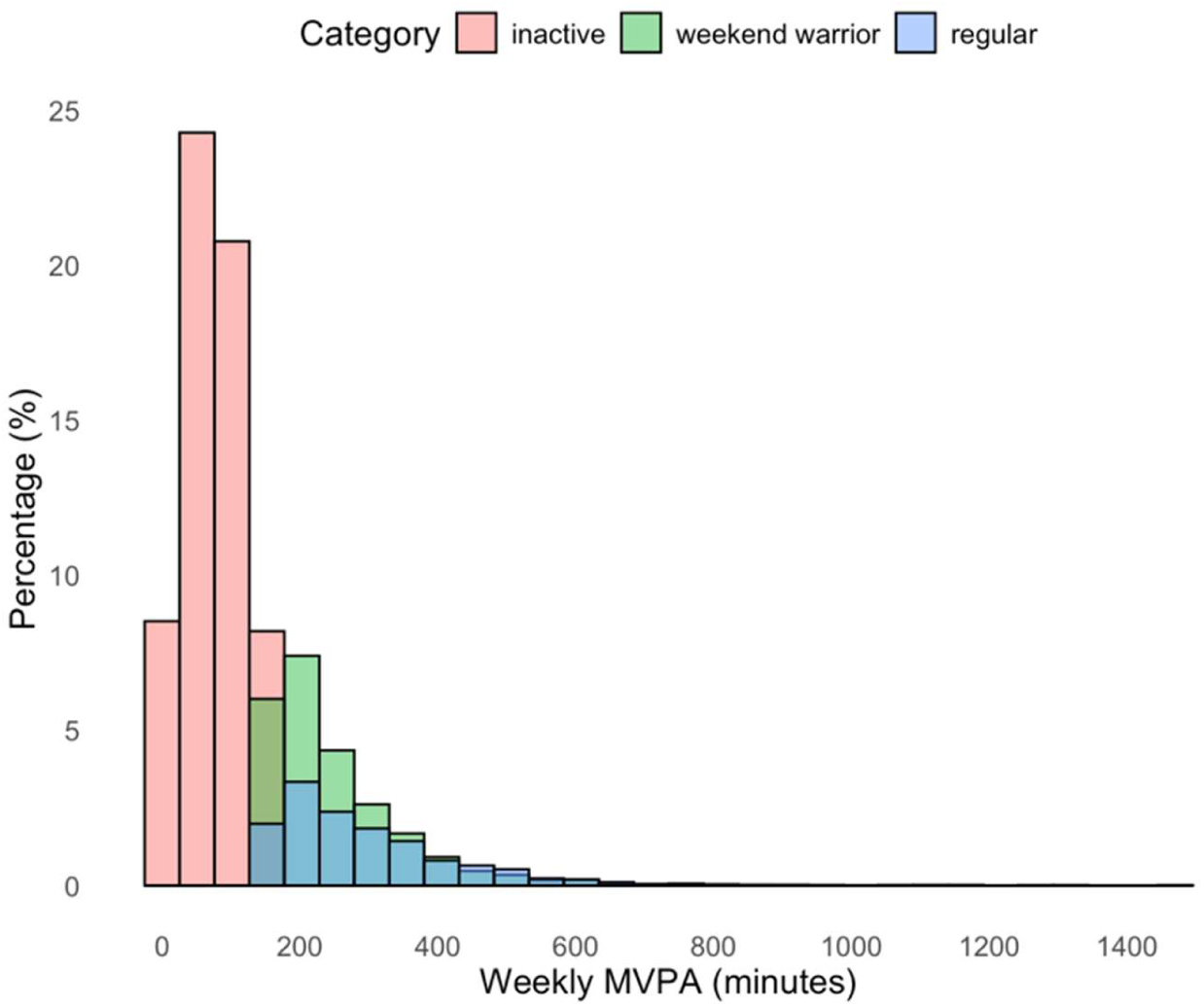
Distribution of MVPA time according to physical activity patterns.

### Associations Between Physical Activity Patterns and Composite Cardiometabolic Health Score

Associations between PA patterns and the composite cardiometabolic health z-score are shown in Figure 2 and Figure S3. Across all models, participants in the WW and regularly active groups had more favorable cardiometabolic health (z-score <0) than inactive participants. In the fully adjusted model, the cardiometabolic health z-score was lower by 0.118 (95% CI, -0.142 to -0.095) in the WW group and by 0.111 (95% CI, -0.141 to -0.081) in the regularly active group, compared with inactive individuals. We found no evidence of a difference between the WW and regularly active groups.

**Figure 2.**
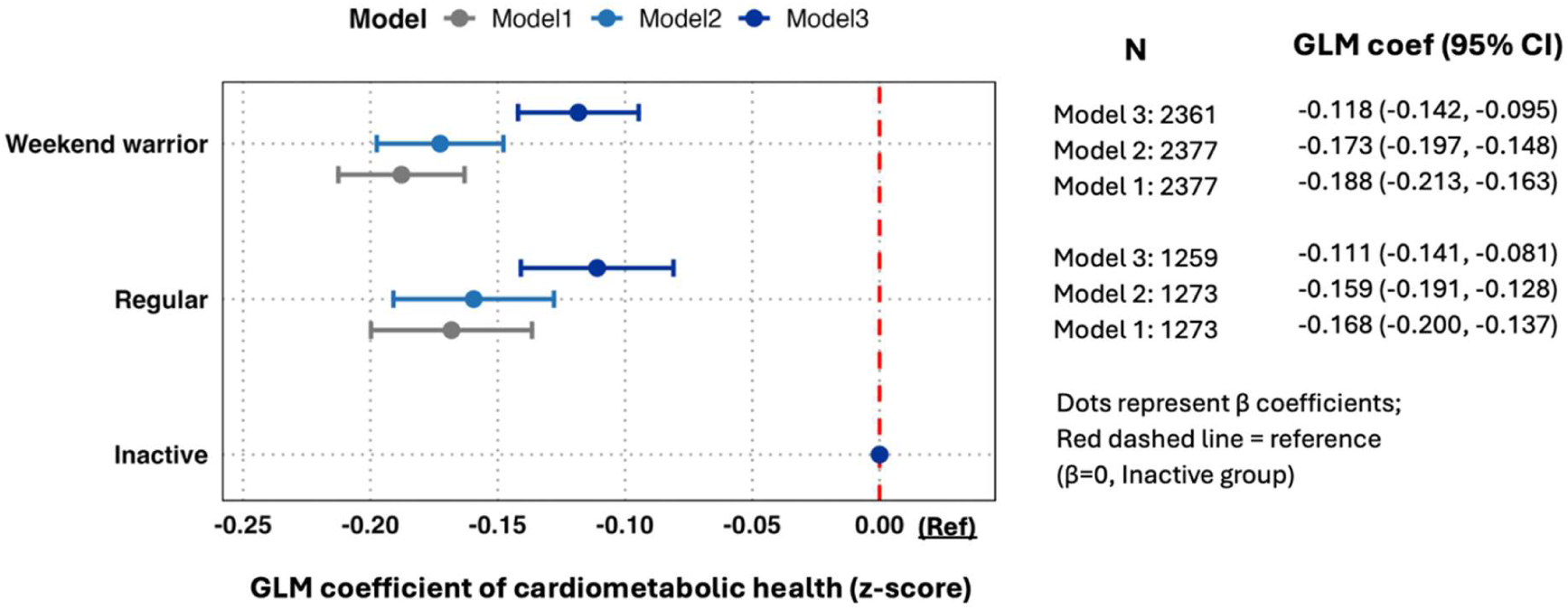
Associations of Physical Activity Patterns and Composite Cardiometabolic Health Score. Model 1 was adjusted for sex, age, and cohort. Model 2 was additionally adjusted for lifestyle behaviors, including alcohol consumption, smoking status, and fasting status (for blood biomarker outcomes). Model 3 was further adjusted for health indicators available across all cohorts: self-rated health, history of CVD, and self-reported use of medications for blood pressure, glucose, and lipid management.

### Associations Between Physical Activity Patterns and Individual Cardiometabolic Health Outcomes

Compared with inactive participants, both WW and regularly active participants showed slightly more favorable profiles across biomarkers, adiposity and blood pressure (Figures 3-4; Model 3 unless stated otherwise).

**Figure 3.**
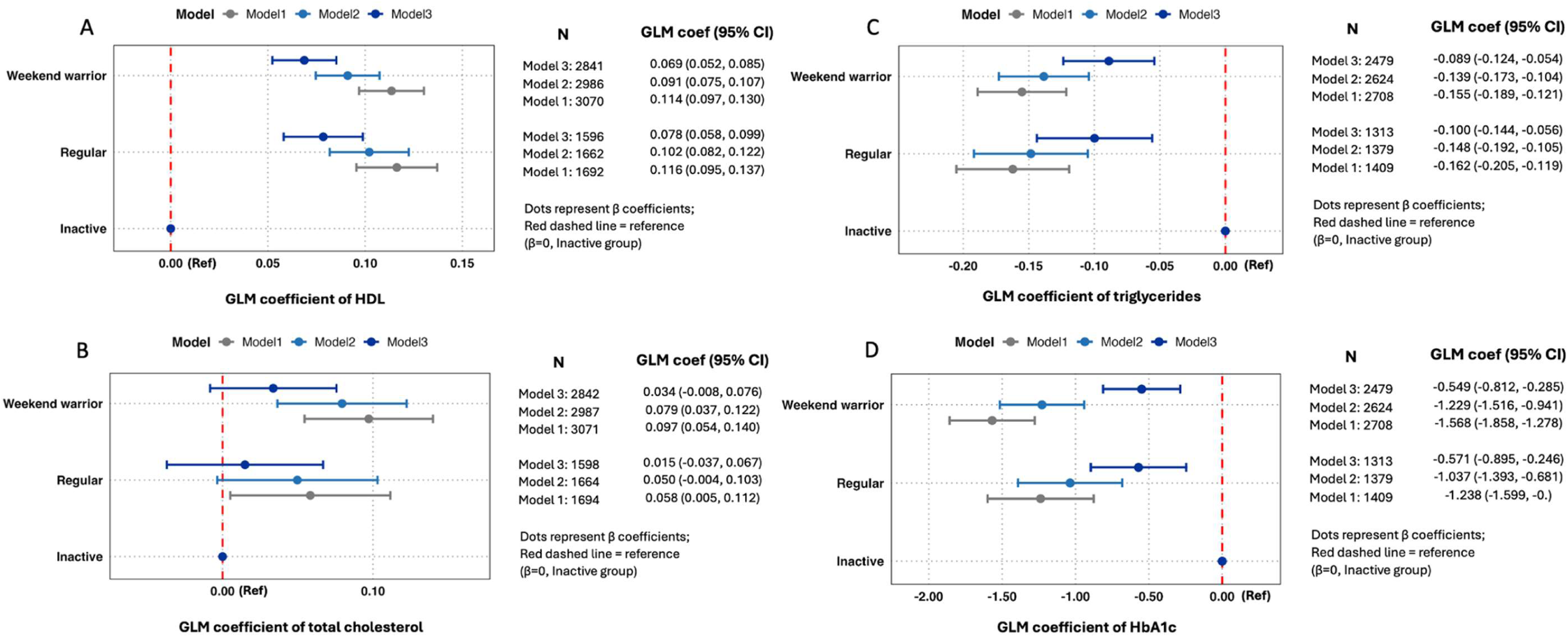
Associations of physical activity patterns with metabolic biomarkers. Adjusted estimates (95% CIs) compare weekend warrior (WW) and regularly active with inactive (<150 min/week MVPA). Outcomes: (A) HDL cholesterol (high-density lipoprotein cholesterol), (B) total cholesterol, (C) triglycerides, and (D) HbA1c (glycated hemoglobin). Active: ≥150 min/week MVPA. Among active, WW: ≥50% of weekly MVPA accrued on the 2 most active days (not required to be weekend/consecutive); others classified as regularly active. Model 1 was adjusted for sex, age, and cohort. Model 2 was additionally adjusted for lifestyle behaviors, including alcohol consumption, smoking status, and fasting status (for blood biomarker outcomes). Model 3 was further adjusted for health indicators available across all cohorts: self-rated health, history of CVD, and self-reported use of medications for blood pressure, glucose, and lipid management.

**Figure 4.**
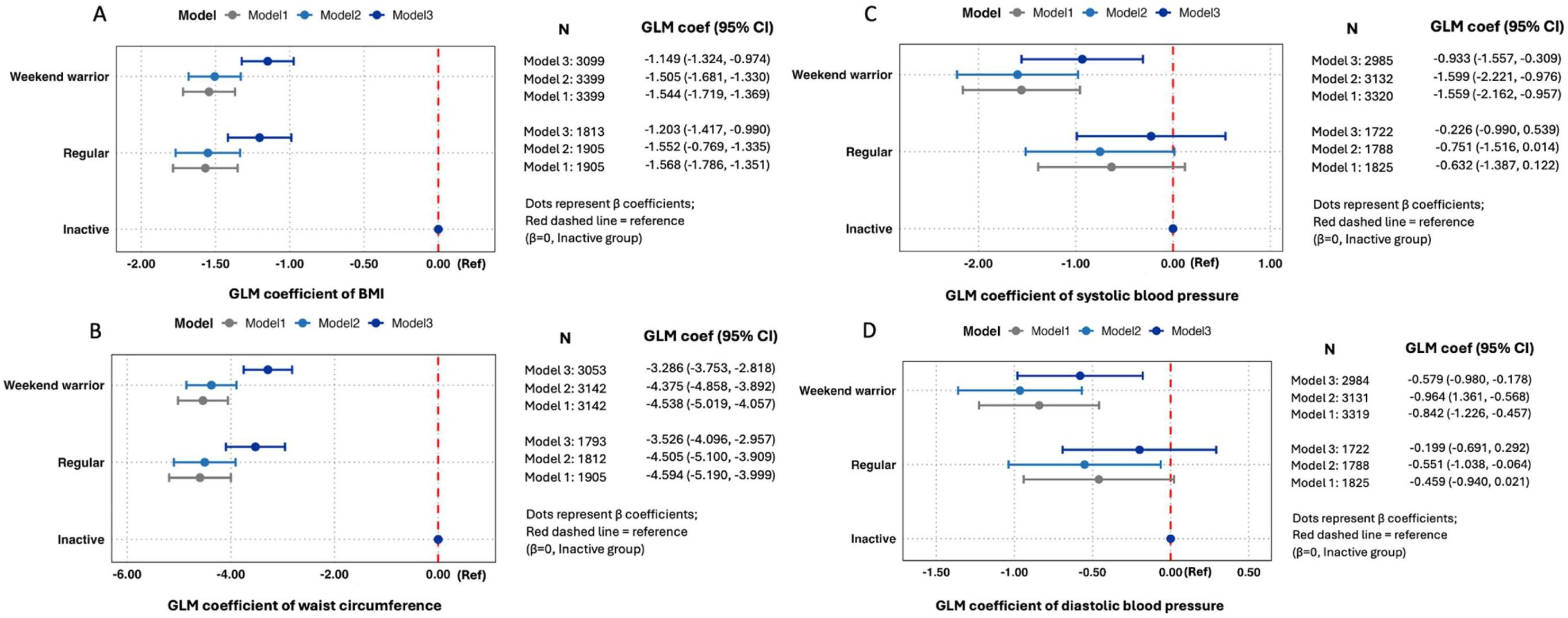
Associations of physical activity patterns with adiposity and blood pressure. Adjusted estimates (95% CIs) compare weekend warrior (WW) and regularly active with inactive (<150 min/week MVPA). Outcomes: (A) body mass index (BMI), (B) waist circumference, (C) systolic blood pressure (SBP), and (D) diastolic blood pressure (DBP). Active: ≥150 min/week MVPA. Among active, WW: ≥50% of weekly MVPA accrued on the 2 most active days (not required to be weekend/consecutive); others classified as regularly active. Model 1 was adjusted for sex, age, and cohort. Model 2 was additionally adjusted for lifestyle behaviors, including alcohol consumption, smoking status, and fasting status (for blood biomarker outcomes). Model 3 was further adjusted for health indicators available across all cohorts: self-rated health, history of CVD, and self-reported use of medications for blood pressure, glucose, and lipid management.

For biomarkers (Figure 3), WW had higher HDL cholesterol (0.069, 0.052-0.085 mmol/L) and lower triglycerides (−0.089, -0.124 to -0.054 mmol/L) and HbA1c (−0.549, -0.812 to -0.285 mmol/mol) than inactive participants; regularly active participants were similar (HDL 0.078, 0.058-0.099 mmol/L; triglycerides -0.100, -0.144 to -0.056 mmol/L; HbA1c -0.571, -0.895 to -0.246 mmol/mol). Associations with total cholesterol were small and not statistically significant after full adjustment (WW 0.034, −0.008 to 0.076 mmol/L; regularly active 0.015, −0.037 to 0.067 mmol/L).

For adiposity and blood pressure (Figure 4), weekend warriors had lower BMI (−1.149, -1.324 to -0.974 kg/m²), waist circumference (−3.286, -3.753 to -2.818 cm), SBP (−0.933, -1.557 to -0.309 mmHg) and DBP (−0.579, -0.980 to -0.178 mmHg) than inactive participants; regularly active participants showed similar adiposity differences (BMI -1.203, -1.417 to -0.990 kg/m²; waist circumference -3.526, -4.096 to -2.957 cm), with blood pressure estimates closer to the null (SBP -0.226, -0.990 to 0.539 mmHg; DBP -0.199, -0.691 to 0.292 mmHg). Weekend warriors did not differ from regularly active adults after full adjustment and in a volume-restricted analysis (Tables S7, S15 and S15a).

### Dose-Response Associations of MVPA with Cardiometabolic Health Score and Outcomes

Multivariable-adjusted dose-response associations between weekly MVPA and the composite cardiometabolic health z-score by activity pattern are shown in Figure 5 (Table S8). Using 150 min/week as the reference, higher MVPA was associated with a lower z-score in both WW and regularly active adults, with overlapping confidence intervals across patterns. The pre-specified nested-model F test provided no evidence of departure from linearity for the z-score (*P* nonlinear = 0.513), and there was no evidence that the MVPA z-score association differed by activity pattern (*P* interaction = 0.479; Table S8). Dose-response curves for individual outcomes showed broadly similar patterns, with little evidence of non-linearity or MVPA×pattern interaction; uncertainty increased at higher MVPA levels where data were sparse (Figures S4-S5; Table S8).

**Figure 5.**
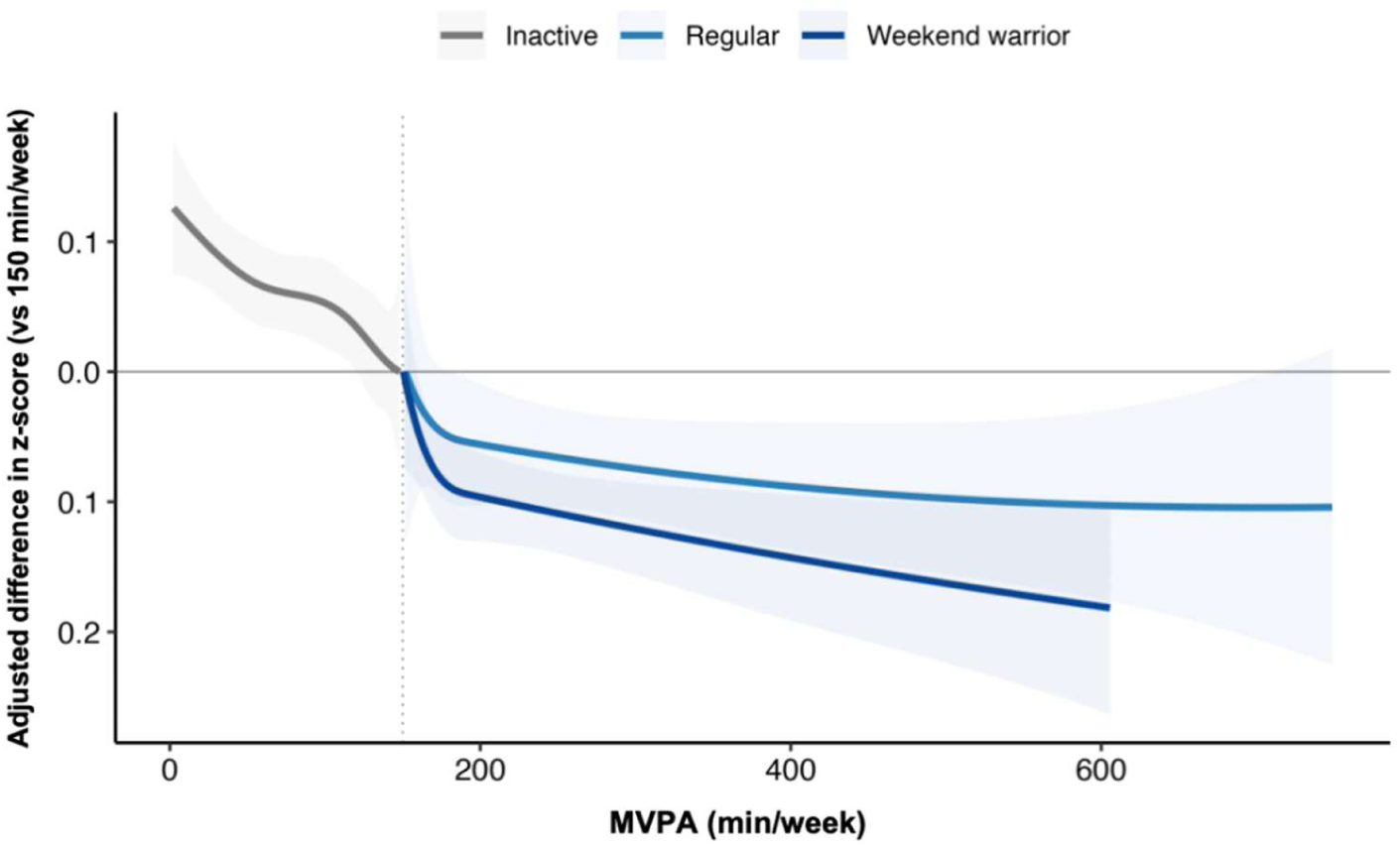
Dose-response association of MVPA with cardiometabolic health score. Multivariable-adjusted dose-response associations between weekly MVPA and cardiometabolic health z-score by PA pattern group, shown as adjusted differences relative to 150 min/week (reference). Shaded areas indicate 95% CIs; histograms show the MVPA distribution within each group. Estimates for the inactive group at/above 150 min/week rely on extrapolation due to sparse data at higher MVPA.

### Alternative Physical Activity Definitions and Sensitivity Analyses Results

Association results were comparable when using an alternative weekend warrior definition (≥75% of weekly MVPA on 1-2 days; Figures S6-S7; Tables S9-S10) and when using the sample median MVPA (≥117.1 min/week) as the activity threshold (Figures S8-S9; Tables S11-S12). Under stricter wear-time criteria (≥5 valid wear days [≥20 h/day] and ≥1 weekend day observed) using the primary (≥50%) WW classification, WW remained statistically similar to regularly active adults (β =-0.004, 95% CI [-0.037, 0.029]), whereas inactive adults had a higher z-score (β=0.099, 95% CI [0.069,0.128]); SBP and DBP differences were not statistically significant (Table S13). Findings were robust after accounting for MVPA volume (additional adjustment and volume restriction). In the volume-restricted analyses, WW and regularly active adults had no statistically significant differences in SBP or DBP, and both groups had lower BP than inactive adults (Tables S14, S15, and S15a).

In sensitivity analyses additionally adjusting for education, occupational class, diet (fruit and vegetable consumption), and mobility limitations, the estimated differences in cardiometabolic health z-score for WW and regularly active adults (vs inactive) were only modestly attenuated (Table S16). Across these models, coefficients for the z-score ranged from -0.117 to -0.099 for weekend warriors and from -0.110 to -0.074 for regularly active adults, compared with inactive adults. For metabolic biomarkers, estimates for HDL cholesterol, total cholesterol, triglycerides, and HbA1c remained similar in magnitude across the extended models. For adiposity and blood pressure, associations with BMI and waist circumference were also largely unchanged, whereas associations with systolic and diastolic blood pressure were small and less consistent.

Additionally, applying a 10-mmHg adjustment for SBP and DBP among participants using antihypertensive medications yielded very similar effect estimates and did not materially alter the conclusions regarding the associations between PA patterns and blood pressure (Table S17). Results were similar in analyses excluding antihypertensive medication users or participants with a history of CVD (Tables S18-21).

## Discussion

In this large pooled IPD analysis of 13,904 adults from six cohorts with device-measured PA, achieving at least the guideline-recommended MVPA volume (≥150 min/week) was associated with a more favorable cardiometabolic profile compared with inactivity, regardless of whether MVPA was concentrated on the two most active days (WW pattern) or not concentrated (regularly active pattern). Both activity patterns were associated with lower adiposity (BMI and waist circumference), higher HDL cholesterol, lower triglycerides, lower HbA1c, and modestly lower systolic and diastolic blood pressure, while no association was observed with total cholesterol. When directly comparing the two active patterns, we observed no meaningful differences for the composite cardiometabolic health score or any individual marker. Dose-response analyses further suggested that, among adults with sufficient MVPA volume (WW and regularly active groups), higher weekly MVPA beyond 150 min/week was generally associated with incremental improvements in the composite score and individual outcomes, with inference limited in the inactive group at higher MVPA levels due to sparse data. These findings were robust across sensitivity analyses using alternative WW definitions and MVPA thresholds, stricter wear-time criteria, and additional covariate adjustment, including models additionally adjusting for total weekly MVPA. Despite statistical significance in this large sample, several effect sizes were small (e.g., HbA1c ∼0.5 mmol/mol and SBP <1 mmHg), and statistical and clinical significance should not be conflated.^23^

Leveraging the pooled ProPASS Consortium,^24^ we applied harmonized definitions of WW and regularly active patterns, a consistent MVPA intensity threshold, and a standardized cardiometabolic health score. Compared with conventional study-level meta-analyses and single-cohort analyses, this IPD approach enables consistent exposure and outcome harmonization across cohorts, reduces between-study heterogeneity due to analytic differences, and improves precision to detect modest differences between activity patterns.^10^ The inclusion of adults from five countries further supports the generalizability of these findings beyond UK^13,15,46^ and US^14,16,47,48^ that previous studies covered. In addition, device-based PA minimizes recall and social desirability biases inherent in self-reported PA.^11,14^

Consistent with the main results, our findings suggest that, within the classification framework, adults who accumulated ≥150 min/week of MVPA had a more favorable cardiometabolic profile than those accumulating <150 min/week, regardless of whether MVPA was concentrated on the two most active days (WW) or not concentrated (regularly active). While previous studies have established that both WW and regular activity patterns are associated with lower risks of cardiovascular and all-cause mortality,^9,11,13,14^ our study extends this evidence to intermediate cardiometabolic markers, and the use of a harmonized composite cardiometabolic score alongside key individual markers provides a broader assessment of cardiometabolic health.^10,11,18,49–51^

Biological mechanisms may help explain why the WW and regularly active patterns were associated with broadly similar cardiometabolic profiles in our analyses. Regular MVPA improves insulin sensitivity, promotes triglyceride clearance, increases HDL cholesterol, and reduces visceral adiposity and inflammation.^52,53^ These adaptations accrue over time and appear to depend primarily on the overall “dose” of activity rather than its daily distribution. Accordingly, concentrating MVPA into 1-2 days may yield comparable intermediate benefits to spreading it across the week.

In contrast, associations with total cholesterol were small and not statistically significant after full adjustment. This pattern is consistent with evidence that PA interventions often produce modest or inconsistent changes in total cholesterol.^54^ Total cholesterol is strongly influenced by diet quality and other lifestyle and metabolic factors (e.g., adiposity/body composition, alcohol intake, and medication use),^55^ which may make it less sensitive to variations in weekly MVPA than HDL cholesterol and triglycerides.

For blood pressure, effect estimates were modest and we observed no clear differences between the weekend warrior and regularly active patterns. These results should be interpreted cautiously because blood pressure is particularly susceptible to attenuation from short-term variability, measurement conditions, and treatment-related factors that are difficult to fully capture (e.g., sodium intake, stress, sleep, and medication timing/adherence), even when overall confounding control is similar across outcomes in observational analyses. In addition, we could not characterise the context of MVPA (leisure, occupational, or transport) or within-day accumulation features; domain-specific activity may have different cardiometabolic correlates (the “PA paradox”).^56^ If regularly active adults accrued a larger occupational component than weekend warriors, this could contribute to weaker blood-pressure associations despite similar weekly MVPA thresholds. In addition, dose-response analyses suggested attenuation toward the null at higher weekly MVPA levels, and regularly active adults accumulated more MVPA on average than WW, which may partly explain modest or inconsistent blood-pressure estimates. Future work integrating domain-specific measures and 24-hour time-use (sleep, sedentary time, and physical activity) is needed to clarify these findings. Although termed “weekend warrior,” our definition reflects concentration of MVPA on the two most active days and does not necessarily correspond to weekend days.

### Implications for Public Health

Our findings suggest that, on average, achieving at least 150 min/week of MVPA was associated with similarly favorable cardiometabolic health whether activity was concentrated in the 1-2 most active days (WW) or distributed more regularly across the week; we found no evidence of a difference between these patterns. Our WW definition reflects concentration on the most active days and does not necessarily mean activity occurred on weekend days; thus, messaging should emphasise achieving the weekly MVPA dose rather than prescribing specific days. Because we did not measure activity domain (e.g., leisure, occupational, transport) or explicitly distinguish weekday versus weekend accumulation, future studies should evaluate whether activity context modifies these associations. As this study focused on the weekly distribution of MVPA, we did not assess whether moderate or vigorous intensity modifies these associations beyond total MVPA volume. A consideration for future studies was MVPA in our study was derived from 1-minute cadence estimates;^36^ prior work has demonstrated that vigorous activity intensity in everyday life typically occurs in bouts shorter than one minute,^6,22,39,57–59^ meaning our approach likely underestimates true VPA volume and precludes reliable intensity-specific analyses. Furthermore, the equivalence between vigorous and moderate PA may not be well captured by a single fixed conversion factor (e.g., 1:2), and evidence suggests that VPA may confer additional cardiometabolic benefits even at low weekly doses. Future studies using higher-resolution accelerometry (e.g., second-by-second epoch data) should examine weekend warrior associations across intensity-specific bands to better characterise the relative contributions of moderate and vigorous activity to the observed health benefits.

### Strengths and Limitations

To our knowledge, this large-scale one-stage pooled IPD analysis is the first to use harmonized individual participant data from thigh-worn accelerometers across six international cohorts. This approach enhances generalizability compared to single-cohort studies and provides precise estimates by applying consistent exposure definitions. Furthermore, device-based measurement eliminates the recall bias inherent in self-reports, while extensive sensitivity analyses confirmed the robustness of our findings.

However, several limitations should be considered. The analyses were cross-sectional, so causality cannot be inferred and reverse causation is possible (i.e., poorer cardiometabolic health may reduce engagement in MVPA), and residual confounding from unmeasured genetic, environmental, or contextual factors may remain despite adjustment. MVPA was assessed over a single week, which may not reflect habitual behavior and could lead to misclassification of activity patterns; additionally, weekend warrior was defined by MVPA concentration on the two most active days rather than specific weekend days. The cohorts were predominantly from high-income settings, potentially limiting generalizability, and the composite cardiometabolic z-score used equal weighting and was not a clinically validated risk score. Finally, because HbA1c was only available in ALSWH, BCS70, and TMS, composite-score analyses were based mainly on these cohorts, which may limit representativeness of the composite-score findings for the full six-cohort ProPASS sample.

## Conclusions

In harmonized individual-participant analyses of six device-based cohorts spanning five countries, we observed broadly comparable cardiometabolic profiles among adults achieving at least 150 minutes/week of MVPA whether accumulated on 1-2 most active days (weekend warrior) or distributed more evenly across the week.

## Supplementary Materials

Supplemental Methods S1&S2, Figure S1-S9, and Table S1-S21.

## Abbreviations and Acronyms

PA: physical activity
MVPA: moderate-to-vigorous physical activity
VPA: vigorous physical activity
WHO: World Health Organization
CVD: cardiovascular disease
WW: weekend warrior
BMI: body mass index
HbA1c: Glycated Hemoglobin
HDL: high-density lipoprotein
LDL: low-density lipoprotein
SBP: systolic blood pressure
DBP: diastolic blood pressure

## Data Availability

Access to data is not available directly from the authors of this manuscript. Access to cohort data may be available by contacting individual cohort and following their specific governance and access requirements.

## Acknowledgments

This study is based on data drawn from six observational studies conducted in the Netherlands, the United Kingdom, Australia, Denmark, and Finland. We extend our gratitude to all the participants who contributed their survey data.

## Ethical Approval

Ethical approval was provided by each individual study during data collection and permitted use of data for secondary analysis (e.g. consortium).

